# Performance of Deep Learning Models in Predicting the Nugent Score to Diagnose Bacterial Vaginosis

**DOI:** 10.1101/2024.09.16.24313614

**Authors:** Naoki Watanabe, Tomohisa Watari, Kenji Akamatsu, Isao Miyatsuka, Yoshihito Otsuka

**Affiliations:** Department of Clinical Laboratory, Kameda Medical Center, Higashi-cho 929, Kamogawa, Chiba, Japan; CarbGeM Inc., 1-5-13 Jinnan, Shibuya-ku, Tokyo, Japan

## Abstract

The Nugent score is a commonly used tool for diagnosing bacterial vaginosis; however, its accuracy depends on the skills of laboratory technicians. We aimed to evaluate the performance of deep learning models in predicting the Nugent score, with the goal of improving diagnostic consistency and accuracy. A total of 1,510 vaginal images collected from a hospital in Japan between 2021 and 2023 were assessed. Each image was annotated by laboratory technicians into one of four categories based on the Nugent score—normal vaginal flora, absence of vaginal flora, altered vaginal flora, or bacterial vaginosis. Deep learning models were developed to predict these categories, and their performance was evaluated by comparing the predicted scores with technician annotations. A high magnification model was further optimized and evaluated using an independent test set of 106 images to assess its performance relative to that of the technicians. The deep learning models demonstrated an accuracy of 84% at low magnification and 89% at high magnification in predicting the Nugent score categories. After optimization, the high magnification model achieved 94% accuracy, surpassing the average 92% accuracy of the technicians. The agreement between deep learning model predictions and technician annotations was 92% for normal vaginal flora, 100% for absence of vaginal flora, 91% for altered vaginal flora, and 100% for bacterial vaginosis. The deep learning models demonstrated accuracy comparable to that of laboratory technicians, which indicates their potential utility in improving the diagnostic accuracy of bacterial vaginosis.

**IMPORTANCE:** Bacterial vaginosis is a global health issue affecting women, causing symptoms such as abnormal vaginal discharge and discomfort. The Nugent score is the standard method for diagnosing bacterial vaginosis and is based on manual interpretation of Gram-stained vaginal smears. However, this method relies on the skill and experience of trained professionals, leading to variability in results and challenges in facilities with limited access to such experts. This poses significant challenges for settings with limited access to experienced technicians. The deep learning models developed in this study predict the Nugent score with high accuracy; thus, they can be used to standardize the diagnosis of bacterial vaginosis, reduce observer variability, and enable reliable diagnosis even in settings without experienced personnel. Although larger scale validation is needed, our results suggest that deep learning models may represent a new approach for the diagnosis of bacterial vaginosis.

## INTRODUCTION

Bacterial vaginosis (BV) is a prevalent vaginal condition characterized by a shift from the normal *Lactobacillus* species to Gardnerella vaginalis and other BV-associated bacteria (1). It affects 23– 29% of women worldwide, with regional variations (2). BV is associated with the risk of sexually transmitted infections, including *Chlamydia trachomatis*, *Trichomonas vaginalis* (3), *Mycoplasma genitalium* (4), human papillomavirus (5), and herpes simplex virus type 2 (6). BV is also associated with preterm birth (7) and neonatal complications (8) in pregnant women.

BV is typically diagnosed using the Amsel’s diagnostic criteria (9) and the Nugent score, which is determined by vaginal Gram staining (10). The Amsel criteria evaluate clinical symptoms and signs (9), whereas the Nugent score, ranging from 0 to 10, reflects the bacterial patterns in vaginal specimens (10). The Nugent score is valued for its low cost, quick turnaround time, and minimal equipment requirements. However, its accuracy varies depending on the skill and experience of the clinician.

Recent advances in deep learning, particularly convolutional neural networks (CNNs) (11), have shown promise for pattern recognition in images and speech, with potential applications in medical image classification. In infectious disease research, CNNs have been used for the automated interpretation of blood culture Gram staining (12) and BV classification (13). Wang et al. developed a CNN model to classify Nugent scores into three categories using high-magnification microscopic images, achieving 82% sensitivity and 97% specificity (13). Despite the potential of CNNs for diagnosing BV, improving their accuracy and automation capabilities remains challenging.

In this study, a CNN model was developed to classify vaginal images into four groups based on the Nugent scoring system. Traditionally, the Nugent score uses three categories, with scores ranging from 4 to 6, typically indicating altered vaginal flora. However, a score of 4 may indicate the absence of vaginal flora rather than their alteration. Given the different microscopic patterns of the altered and absent vaginal flora, we refined our model to accurately differentiate between these conditions. We evaluated the proposed BV models using both low- and high-magnification images. Low-magnification images that do not require oil immersion simplify the process and facilitate automation.

## RESULTS

### Prediction performance of the BV model

Table 1 shows the agreement between the predicted classifications of the BV model and true label groups. The high-magnification model accurately predicted 277 of 310 samples based on the Nugent score, whereas the low-magnification model identified the correct category in 260 of the 310 samples. Table 2 presents the agreement and accuracy rates for both high- and low-magnification models. In the four-group classification, the high-magnification model demonstrated better agreement rates across all categories. The lowest agreement rate was observed for identifying altered vaginal flora, with the high-magnification model at 57% and the low-magnification model at 50%. In this classification, the high-magnification model achieved an accuracy of 89%, surpassing that of the low-magnification model (84%).

**TABLE 1.** Estimated group of BV models for true label.

| True label | No. of samples, low-magnification model |  |  |  | No. of samples, high-magnification model |  |  |  |
| --- | --- | --- | --- | --- | --- | --- | --- | --- |
|  | Normal | No flora | Altered | BV | Normal | No flora | Altered | BV |
| Normal (n = 70) | 70 | 0 | 0 | 0 | 56 | 0 | 14 | 0 |
| No flora (n = 60) | 0 | 60 | 0 | 0 | 0 | 59 | 0 | 1 |
| Altered (n = 40) | 8 | 0 | 12 | 20 | 0 | 0 | 38 | 2 |
| BV (n = 140) | 0 | 10 | 12 | 118 | 0 | 1 | 15 | 124 |

**Footnotes:** Normal, normal vaginal flora; No flora, no vaginal flora; Altered, altered vaginal flora; BV, bacterial vaginosis.

**TABLE 2.** BV prediction comparison of low and high-magnification models.

| Evaluation index | Performance of CNN model |  |
| --- | --- | --- |
|  | low-magnification model | high-magnification model |
| Agreement rates in four-group classifications (%) |  |  |
| Normal vaginal flora (n = 70) | 90 | 100 |
| No vaginal flora (n = 60) | 86 | 98 |
| Altered vaginal flora (n = 40) | 50 | 57 |
| Bacterial vaginosis (n = 140) | 86 | 98 |
| Agreement rates in two-group classifications (%) |  |  |
| Bacterial vaginosis (n = 180) | 100 | 92 |
| No bacterial vaginosis (n = 130) | 88 | 99 |
| Accuracy, (%) |  |  |
| Four-group classifications | 84 | 89 |
| Two-group classifications | 94 | 95 |

**Description:** The agreement rate is defined as the percentage of results from the CNN model that matches the true label.

Of the 310 samples, 130 were classified as non-BV and the remaining 180 were classified as BV. In the two-group classification, the low-magnification model had an accuracy of 94% (292/310), which was slightly lower than that of the high-magnification model (95%, 294/310). For the BV group, the agreement rate with the 400× model reached 100%, which was higher than that of the high-magnification model (92%). In the non-BV group, the agreement rate was lower (88%) for the low-magnification model than that of the high-magnification model (99%).

### Development and provisional performance of the advanced BV model

The high-magnification model, which initially exhibited greater accuracy, was further improved through additional learning. For this purpose, 430 new images were included for a total of 1,510 images used to develop the advanced BV model. The revised image distribution across the Nugent score categories included 450 images of normal vaginal flora, 490 images of no vaginal flora, 300 images of altered vaginal flora, and 700 images of bacterial vaginosis. In the interim evaluation, the advanced BV model achieved an accuracy rate of 92% in the four-group classification, representing a 3% improvement over an earlier version of the model.

### Comparison of the advanced BV model and human experts in predicting BV

To assess the performance of the advanced BV model in differentiating between bacterial vaginosis and non-BV cases, an image was obtained from each of the 106 vaginal discharge specimens. The composition of these samples was as follows: 61 (58%) had normal vaginal flora, 10 (9%) had no vaginal flora, 14 (13%) had altered vaginal flora, and 21 (20%) had BV. These were classified into 71 non-BV (67%) and 35 BV (33%) samples. Table 3 shows the agreement between the predicted classifications of the advanced BV model and true label groups. For four-group classification, the advanced BV model achieved an accuracy of 94% (Table 4). The accuracies observed for the two laboratory technicians were 87% and 96%, respectively, and the collective average accuracy for the laboratory technicians was 92%. Altered vaginal flora had the lowest prediction accuracy, whereas the advanced BV model showed a 91% agreement rate.

**TABLE 3.** Prediction performance of the advanced BV model.

| Model or technician | Group | True label |  |  |  |
| --- | --- | --- | --- | --- | --- |
|  |  | Normal<br>(n = 61) | No flora<br>(n = 10) | Altered<br>(n = 14) | BV<br>(n = 21) |
| Advanced BV model | Normal | 61 | 0 | 4 | 1 |
|  | No flora | 0 | 10 | 0 | 0 |
|  | Altered | 0 | 0 | 10 | 1 |
|  | BV | 0 | 0 | 0 | 19 |
| Technician 1 | Normal | 61 | 0 | 2 | 5 |
|  | No flora | 0 | 10 | 0 | 0 |
|  | Altered | 0 | 0 | 11 | 6 |
|  | BV | 0 | 0 | 1 | 10 |
| Technician 2 | Normal | 58 | 0 | 0 | 0 |
|  | No flora | 0 | 10 | 0 | 0 |
|  | Altered | 3 | 0 | 13 | 0 |
|  | BV | 0 | 0 | 1 | 21 |

**Footnotes:** Normal, normal vaginal flora; No flora, no vaginal flora; Altered, altered vaginal flora; BV, bacterial vaginosis.

**TABLE 4.** Prediction comparison between the advanced BV model and human experts.

| Group and evaluation index | Advanced BV model | Technician average | Technician 1 | Technician 2 |
| --- | --- | --- | --- | --- |
| Agreement rates in four groups (%) |  |  |  |  |
| Normal vaginal flora (n = 61) | 92 | 95 | 90 | 100 |
| No vaginal flora (n = 10) | 100 | 100 | 100 | 100 |
| Altered vaginal flora (n = 14) | 91 | 73 | 65 | 81 |
| Bacterial vaginosis (n = 21) | 100 | 93 | 91 | 95 |
| Agreement rates in two groups (%) |  |  |  |  |
| Bacterial vaginosis (n = 35) | 100 | 96 | 100 | 92 |
| No bacterial vaginosis (n = 71) | 93 | 96 | 91 | 100 |
| Accuracy |  |  |  |  |
| Four group (%) | 94 | 92 | 87 | 96 |
| Two group (%; 95% CI) | 95 (89–99) | 95 (NA) | 93 (87–97) | 97 (92–99) |
| Sensitivity (%; 95% CI) | 86 (70–95) | 90 (NA) | 80 (63–92) | 100 (86–100) |
| Specificity (%; 95% CI) | 100 (93–100) | 98 (NA) | 100 (93–100) | 96 (88–99) |

**Footnotes:** NA, not applicable; Technician, laboratory technician.

In the two-group classification, both the advanced BV model and technicians demonstrated sensitivities greater than 80%, specificities greater than 96%, and accuracies greater than 93%. The sensitivity of the advanced BV model was 86% (95% CI: 70–95%), which was 4% lower than the average sensitivity of 90% achieved by the technicians. Conversely, the specificity of the advanced BV model was 100% (95% CI: 93–100%), which was 2% higher than that of the technicians. The overall accuracy of the advanced BV model was 95% (95% CI: 89–99%), which was comparable to the average accuracy reported by the technicians. Among the BV predictions, 14% (5/35) of the samples identified as BV were incorrectly classified as non-BV by the advanced model, of which four were classified as altered vaginal flora and one was classified as BV.

### Agreement level between the advanced BV model and laboratory technicians

The advanced BV model achieved an overall agreement rate of 92% (98 out of 106) with both laboratory technicians. The kappa coefficient indicated an almost perfect agreement of 0.81 (range 0.68–0.94) between the advanced BV model and technician 1, and an almost perfect agreement of 0.83 (range 0.71–0.94) with technician 2. The inter-technician agreement rate was 91% (96 out of 106), with a kappa coefficient of 0.78 (range 0.65–0.91), indicating substantial agreement between technician 1 and technician 2.

## DISCUSSION

We developed a CNN model to predict Nugent scores from vaginal Gram stains and achieved 94% accuracy across a four-group classification. This result surpassed the performance reported by Wang et al. (13), who achieved 80% accuracy for three Nugent score groups in a test set created from images at a single facility. Our CNN model differs from that proposed by Wang et al. with respect to the underlying base model, which includes an additional Nugent score group. Our approach used ConvNeXt (14), which differed from the EfficientNet (15) used by Wang et al. (13). Further, their model categorized scores into three groups, whereas our study expands these to four groups. These changes likely contributed to the improved model accuracy.

Our model effectively matched the laboratory technicians in classifying BV and non-BV with an accuracy of 95%, sensitivity of 86%, and specificity of 100% in the two-group classification. Wang et al. reported a sensitivity of 89% and a specificity of 85% (13). Although our model showed sensitivities <90%, similar to the model by Wang et al., it primarily misclassified samples with altered vaginal flora as normal flora. Moreover, both the CNN models and human technicians found it difficult to accurately identify altered vaginal flora, as evidenced by the low average agreement rate of 73%. Therefore, the accuracy of the CNN model must be improved, particularly for samples with altered vaginal flora.

A significant advantage of low-magnification images is their compatibility with automated microscopy platforms, which simplifies image acquisition. Smith et al. used an automated microscopy platform for collecting Gram-stained images at 400× magnification to develop a CNN model (12). In our study, although the low-magnification model achieved 94% accuracy in the two-group classification, it only achieved 84% accuracy in the four-group classification, highlighting the limitations of using low-magnification images in automated BV scoring. Future improvements, including refining the model by integrating more accurately classified samples, are thus crucial to improve the reliability of automated BV scoring.

BV is a common condition in women, typically diagnosed using conventional methods and nucleic acid amplification tests (NAATs) (16–18). Conventional diagnostic tools include the Nugent score (10), Amsel’s diagnostic criteria (9), OSOM BV Blue assay (19, 20), and FemExam card (21). NAATs, such as the BD Max vaginal panel (22) and Hologic Aptima BV (23) are also used. The Nugent score, which is often used as a reference method, demonstrates substantial inter-observer agreement with kappa coefficients ranging from 0.70 to 0.77 (24) and inter-center agreement ranging from 0.60 to 0.72 (25). However, interpretation of the Nugent score requires expertise, which affects its reproducibility. Our CNN model shows high BV prediction performance and provides results independent of technician skill and subjectivity, with excellent agreement rates (kappa coefficients of 0.81–0.83 with technicians). Implementing this CNN model in a clinical setting could facilitate objective and reproducible interpretation of vaginal Gram staining; hence, aiding in BV diagnosis.

This study has some limitations, particularly in terms of generalizability and sample size. The evaluation was limited to a single institution, which may have limited the broader applicability of the results. Factors such as sample diversity, variations in image hue, and technician skills, which may vary among institutions, could affect the model accuracy. Furthermore, the CNN model was developed using a relatively modest dataset of less than 2,000 samples, which may result in undertraining and affect predictive ability. Despite these limitations, our CNN model demonstrated sensitivity, specificity, and accuracy comparable to those of technicians in the two-group classification. With an expanded dataset, we anticipate significant improvements in the predictive performance of the model, further refining its effectiveness for BV diagnosis when tested on a broader range of samples and settings.

In conclusion, we developed a CNN model to automatically predict BV scores, achieving an accuracy rate of 94% in the four-group classification using high magnification images. These results highlight the potential of CNN models for future applications in the automated classification of BV scores. Currently, there are limited data on the use of CNN models to predict BV scores. To establish its efficacy, this CNN model requires further validation using different vaginal specimens and clinical settings.

## MATERIALS AND METHODS

This study was conducted at the Kameda Medical Center in Japan from November 2021 to February 2024. Figure 1 shows the flowchart of this study. After data collection and preprocessing, two magnification versions of the CNN model were developed for comparative evaluation. The more effective model of these was subsequently selected, improved, and subjected to final evaluation. Ethical approval was obtained from the Kameda Medical Center Ethics Committee (approval number 22-128). The requirement for written informed consent from the participants was waived by the Research Ethics Committee because of the exclusive use of anonymized data in this study.

**Figure 1.**
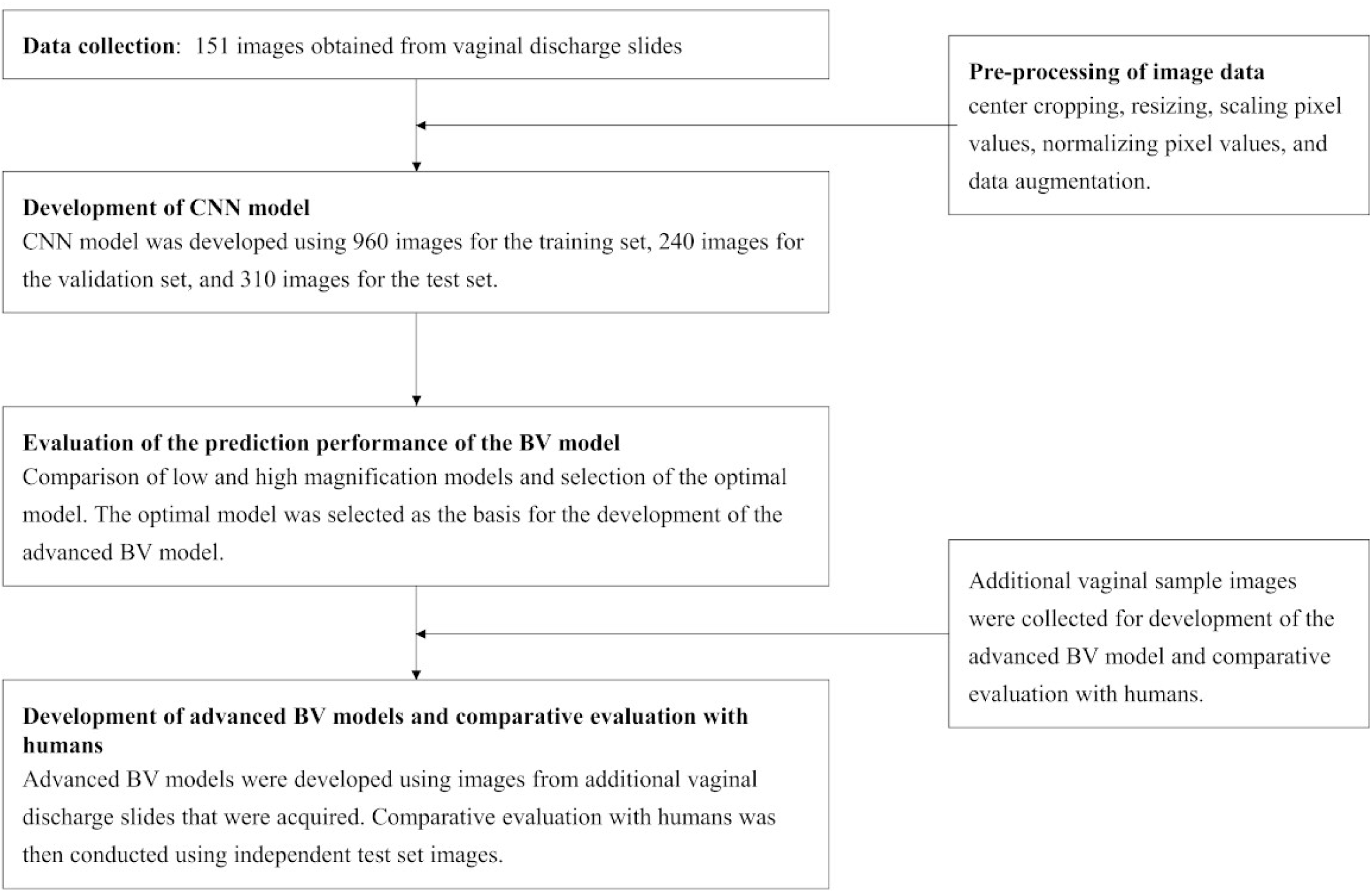
Flowchart of the bacterial vaginosis model development and evaluation.

### Data collection

From November 2021 to May 2023, we collected 151 Gram-stained slides from 151 vaginal discharge specimens. Gram staining was performed using Neo-B & M Wako crystal violet solution, iodine solution, decolorizing solution, and Pfeifel solution (FUJIFILM Wako Chemicals, Osaka, Japan). A Nikon ECLIPSE Ci-S microscope equipped with a DS-Fi3 digital camera was used for image acquisition. The images, focused on areas where bacteria or cells were visible, were captured at 400× (low) and 1,000× (high) magnification, each with a resolution of 2,880 × 2,048 pixels.

Images were categorized into four groups according to the Nugent score: normal vaginal flora (score 0–3); no vaginal flora (score 4), altered vaginal flora (scores 5–6); or BV (score 7–10). Figure 2 shows the representative slide images for each group. Nugent scores were assessed by two laboratory technicians, including at least one certified clinical microbiology specialist. In cases of disagreement, a third technician was consulted for the final decision. In total, 1,510 images at both low and high magnifications were collected from each slide. Initially, images of BV were collected and based on the Nugent scores, the distribution was as follows: 320 images for normal vaginal flora, 300 for no vaginal flora, 190 for altered vaginal flora, and 700 for BV. These images were randomly allocated to the training, validation, and testing sets with 960, 240, and 310 images, respectively.

**Figure 2.**
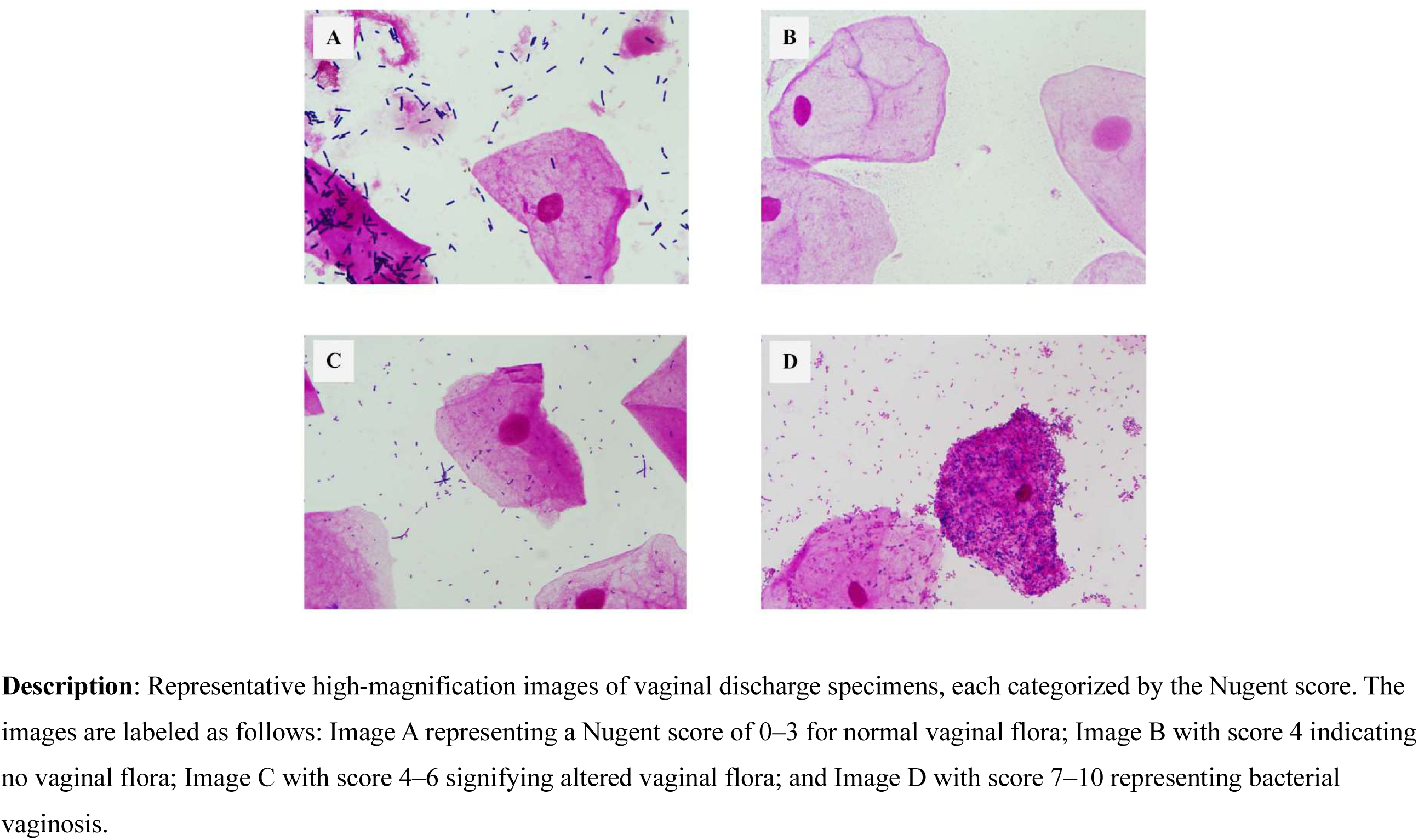
Microscopic images of vaginal discharge specimens and the Nugent Score categories.

### Pre-processing of images and data augmentation

We applied four preprocessing steps to the collected images: center cropping, resizing, scaling pixel values, and normalizing pixel values. Microscopic images were cropped from their original size of 2,880 × 2,048 pixels to a central area of 2,048 × 2,048 pixels. The cropped images were resized to 1,024 × 1,024 pixels. The pixel values were scaled from (0, 255) to (0, 1) and normalized to RGB means of (0.485, 0.456, and 0.406) and RGB standard deviations of (0.229, 0.224, and 0.225).

To improve the model performance, data augmentation techniques were implemented during the learning process. These techniques included random rotation, random cropping, random horizontal and vertical flipping, random affine transformations, and color jittering. Random rotation and cropping involved arbitrary rotations and adjustments of image dimensions. Random horizontal and vertical flipping altered images by flipping them left/right and up/down, respectively. Random affine transformations and color jittering variably adjusted the affine parameters of brightness, contrast, saturation, and hue.

### Development of the BV model using a CNN

Neural networks are mathematical models that emulate the functions of nerve cells in the human brain. Specifically, in image classification, these networks learn to recognize image content by iteratively processing the training data, thereby updating the connections between neurons. Among the various types of neural networks, CNNs are tailored to process image data. In our study, we used a model based on ConvNeXt, a variant of a CNN known for its state-of-the-art performance in image classification, including its high accuracy and scalability (14). We used a linear activation function in the final layer of the BV model to compute the probabilities representing the likelihood of each Nugent score group. This step is essential for effectively predicting Nugent scores based on the analyzed images.

### Evaluation of the prediction performance of the BV model

The predictive performance of the BV model was evaluated for both the four- and two-group classifications derived from the BV categories. For the two-group classification, the four Nugent scores were divided into two categories: BV and non-BV, with normal and no vaginal flora being categorized as non-BV whereas altered vaginal flora and bacterial vaginosis were categorized as BV.

We used agreement rate and accuracy as the evaluation metrics. The agreement rate measures the consistency between the CNN model predictions and the actual labels and is expressed as a percentage. Accuracy is the proportion of correct predictions made by the CNN model compared with the actual labels over the entire dataset. For the two-group classifications, sensitivity and specificity were calculated as follows: sensitivity was the ratio of correctly predicted BV cases to the total number of actual BV cases; and specificity was the ratio of correctly predicted non-BV cases to the total number of actual non-BV cases.

### Development of an advanced BV model

Among the models developed using low- and high-magnification images, the model with superior accuracy in the four-group classification was selected for further refinement. This refinement process included the integration of additional images collected between August and October 2023, using the same methodology as in the initial development phase. We applied RandAugment (26), a method used to simplify and improve data augmentation techniques. The performance of this advanced BV model was assessed on an interim basis using the same test set of 310 images used in the initial evaluation.

### Accuracy comparison between the advanced BV model and human assessment in BV diagnosis

An independent test set was used to compare the accuracy of the advanced BV model with that of human experts. An image was acquired for each vaginal discharge specimen collected in December 2023. These images were labeled based on the criteria established during BV model development. These data were used to evaluate and compare the agreement rate, accuracy, and kappa coefficients between the advanced BV model and the laboratory technicians. Kappa coefficients were calculated to evaluate agreement between the advanced BV model and the laboratory technician. Statistical analyses were conducted using EZR version 1.64 (27).

## Data Availability

All the data supporting the findings are provided in the article. Any additional data are available on request to the corresponding authors.

## DATA AVAILABILITY

All the data supporting the findings are provided in the article.

## ACKNOWLEDGMENTS

We thank the technicians at Kameda Medical Center for their assistance with vaginal specimen collection and data handling.

